# Reduced *Bacille Calmette-Guérin*-specific IgG titres among babies born to mothers with Active Tuberculosis Disease in Uganda

**DOI:** 10.1101/2024.10.11.24315362

**Authors:** Diana Sitenda, Phillip Ssekamatte, Rose Nakavuma, Andrew Peter Kyazze, Felix Bongomin, Joseph Baluku, Rose Nabatanzi, Davis Kibirige, Annette Nakimuli, Stephen Cose, Irene Andia-Biraro

## Abstract

**Background:** Babies born to mothers with active tuberculosis disease (ATB) are at risk of poor clinical outcomes such as low birth weight and perinatal mortality. However, little is known about the influence of maternal ATB exposure on their vaccine responses during infancy. The study aimed to explore how maternal ATB affects infants’ vaccine responses, hypothesising reduced responses to BCG and other infant vaccines.

**Methods:** This was a case-control study with a longitudinal component of babies born to mothers with bacteriologically confirmed ATB (cases) and babies born to mothers without ATB (controls) carried out between September 2021 and June 2022. Quantitative BCG, diphtheria, tetanus, and measles-specific IgG ELISA assays were performed on infant plasma harvested from lithium-heparin blood collected on first encounter after birth (0), at 3, 6, and 9 months. We used prism v10.1.2, Mixed-effects modelling, and Tukey’s multiple comparison testing to determine mean differences (MD) between the cases and controls at all time points.

**Results:** Infants cases had reduced IgG titres to BCG at baseline compared to the controls (p=0.04), with a mean of 125.8/141.1 IU/mL, respectively. This difference was, however, not sustained at the other time points. Similarly, we demonstrated strong trends of reduced responses to tetanus, diphtheria, and measles vaccines among infant cases at baseline and three-month time points and weakly at months six and nine. The mean titres for tetanus at baseline and 3 months for cases versus controls are 1.744/2.917 IU/mL and 1.716/2.344 IU/mL (p<0.0001/0.018), respectively. The mean titres for diphtheria at 3 months were 0.022/0.075 IU/mL (p=0.006), respectively.

**Conclusion:** We have demonstrated that maternal TB disease influences vaccine responses to BCG and other infant vaccines. This has implications for increased risk of childhood TB and other preventable diseases.

## Introduction

The incidence of Tuberculosis (TB) disease among people aged below 30 years of age has increased, hence increasing the number of childbearing women contracting the disease (1). The TB burden in pregnancy globally is estimated to be 216,500 cases annually, with an increased threefold risk of TB in this population (2). Mother-to-child transmission of TB may occur *in utero* through hematogenous spread via the umbilical vein and aspiration or swallowing of infected amniotic fluid (3, 4). TB disease in pregnancy leads to poor maternal and foetal outcomes such as preterm birth, low birth weight (5), perinatal death, congenital anomalies, small for gestational age, foetal distress, low Apgar scores, and rare congenital tuberculosis (6). There is limited data on the immune responses to BCG and other vaccines of infants born to mothers with active TB (ATB). Vaccination with BCG has been reported to be associated with reduced infant mortality (7, 8) although it still remains unclear whether this is true to all populations including babies born to mothers with active tuberculosis. A study by Lubyayi et al., 2020 revealed that maternal Latent TB status does not affect the infants’ response to BCG-vaccine (9). On the other hand, Mawa et al., 2015 reported a decrease in PPD-specific responses among infants born to mothers with latent tuberculosis infection at one week and six-weeks following vaccination with BCG compared to the control group (10). Both studies explored the impact of maternal latent TB on infant vaccine responses; however, in this study, we assessed vaccine responses among infants born to active TB mothers and compared them with babies born to TB-uninfected mothers. We found it important also to determine if exposure to TB disease influenced the other infant vaccines to pave the way for possible public health-related interventions, given that HIV alone has been implicated in causing a reduced response to vaccines among infants (11).

This study determined the IgG titer responses to *Bacille Calmette-Guérin* (BCG), diphtheria, tetanus, and measles vaccines in babies born to mothers with and without active TB disease at baseline, three, six, and nine months of follow-up.

## Materials and Methods

### Study design and population

This was a case-control study with a longitudinal component. The study enrolled 35 women with bacteriologically confirmed active TB as cases and 33 women without TB as controls matched for maternal and gestational age from antenatal (ANC) and postnatal (PNC) clinics at three major health facilities in Kampala, Uganda: Kasangati Health Center IV, Kisenyi Health Center IV, and Kawempe National Referral Hospital. However, we considered 25 cases (babies born to mothers with active tuberculosis disease) and 25 controls (babies born to mothers without active tuberculosis disease) in our experiments, whose numbers also dropped in successive follow-up months. Upon delivery, babies were assented to, enrolled in the study, and were followed up for nine months. These babies donated up to six milliliters of blood in lithium-heparin tubes at different time points, from which plasma was harvested and stored at −20 in a freezer at the Immunology laboratory of Makerere University College of Health Sciences (MakCHS). The baseline visit (V0) marked the visit of the first encounter with infants aged less than a month (cases, n=19 versus controls, n=15). The V1 marked a visit at three months (cases n=15 versus controls n=14), V2 at six months (cases n=12 versus controls n=8), and V3 at 9 months (cases n=8 versus controls n=4). Vaccine schedules included BCG at birth; pentavalent tetanus-diphtheria-pertusis-hepatitis B-haemophilus influence type B (DPT-HEPB-HIB1) at 6, 10, 14 weeks; and measles at nine months.

### Laboratory methods

Heparin blood was layered over Ficoll-paque in a ratio of 1:2 under a biosafety cabinet in a 15ml Falcon tube. This was spun in a centrifuge at 1800rpm for 30 minutes to separate plasma and PBMCs from red blood cells. Plasma was harvested using a pipette and was stored at −20°C freezer. All assays for each vaccine were optimised to achieve a suitable dilution factor. Archived available plasma samples were retrieved from a −20 C freezer and transferred to the fridge at the temperature of 4 overnight. They were then retrieved from the refrigerator and thawed with the ELISA kits at room temperature (25) for 1 hour. Plasma samples were vortexed to achieve uniform mixing, and 50μl were loaded into designated wells, in addition to standards and blanks, of the 96-well ELISA plates. The human IgG-specific ELISA assays were performed to determine plasma IgG titres of BCG, Diphtheria-Tetanus, and Measles vaccine responses using the human BCG (Cat No. MBS702330), diphtheria (Cat No. MBS495632), tetanus (Cat No. MBS9907877) and measles (Cat No. MBS283708) IgG ELISA kits following the manufacturer’s instructions. The absorbance at 450/620 nm was measured using the Gen 5 software version 2.71.2 for the ELISA reader (Synergy LX multimode reader). Detailed procedures are available in the supplementary materials.

### Data analysis and management

Optical densities (ODs) from the ELISAs were converted to concentrations using the Gen 5 software version 2.71.2 and then exported to Excel. Using Excel, the data was cleaned and grouped as cases versus controls at baseline, 3, 6, and 9 months. Data was statistically analysed in GraphPad Prism v9.5.1 using the Mixed model effect analysis with Tukey’s multiple comparisons test. Means, mean difference (MD), and standard error of difference (SE) for both cases and controls were computed at all time points. A simple linear regression analysis was performed on the relationship between birth weight, haemoglobin, and vaccine responses at all time points. The Mann-Whitney U-test was used to determine statistical differences in baby weights, age, and the mother’s haemoglobin concentrations among both cases and controls. The chi-square test was also used to determine statistical differences in the gender of babies, their postnatal versus antenatal enrolments, and their maternal HIV status.

### Ethical considerations

This study received ethical approval from the School of Medicine Research and Ethics Committee (SOMREC reference number SOM 2020-11); the School of Biomedical Sciences (SOBSREC reference number SBS-2022-226) at Makerere University; and the Uganda National Council for Science and Technology (UNCST registration number HS1396ES). The mothers provided written informed consent and assent for their babies to participate in the study.

## Results

### Characteristics of the study participants

Babies born to mothers with ATB had lower birth weights than those born to mothers without ATB (p=0.03), as shown in Table 1. In addition, case mothers had lower haemoglobin concentrations compared to controls. The median (g/dl) and IQR cases versus controls are 11.2 (10.0-11.9)/ 13.0 (10.7-13.8) (p=0.033). Other characteristics such as age, gender, maternal HIV status, and postnatal versus antenatal babies were found to be similar for both cases and controls.

**Table 1.** shows baseline participant characteristics.

| Babies | Cases (n=25) | Controls (n=25) | P value |
| --- | --- | --- | --- |
| *Weight (Kgs)<br>[median (IQR)] | 3.0 (2.1 - 3.2) | 3.4 (2.7 - 4.4) | 0.03 |
| Age (weeks)<br>[(median (IQR)] | 7 (2-12) | 12 (1-16) | 0.45 |
| Sex (male)<br>(female) | 8<br>12 | 8<br>12 | >0.99 |
| HIV status (positive)<br>(negative) | 7<br>18 | 8<br>13 | 0.47 |
| Antenatal care (ANC)<br>Postnatal care (PNC) | 6<br>19 | 4<br>18 | 0.63 |

\*Means a p-value less than 0.05. Missing values for weight, age, sex, haemoglobin concentration, HIV status, and Antenatal versus postnatal care are 11, 12, 10, 10, 4, and 3, respectively.

### BCG-specific IgG responses

The IgG titres were determined in plasma samples of the case babies versus the control babies. The samples were collected at V0, V1, V2, and V3, and the titres were compared at all time points. The IgG titres were strongly decreased in cases versus controls (p=0.032) at V0. The mean for cases versus controls (IU/mL), mean difference (MD) 95% confidence interval (CI), P-value, and Standard Error (SE)] are shown in Table 2. This was followed by a gradual reduction at the rest of the time points: 3, 6, and 9 months (p=0.574/0.626/0.717), respectively. These differences were, however, not supported, as demonstrated in Figure 1 (A and B).

**Figure 1.**
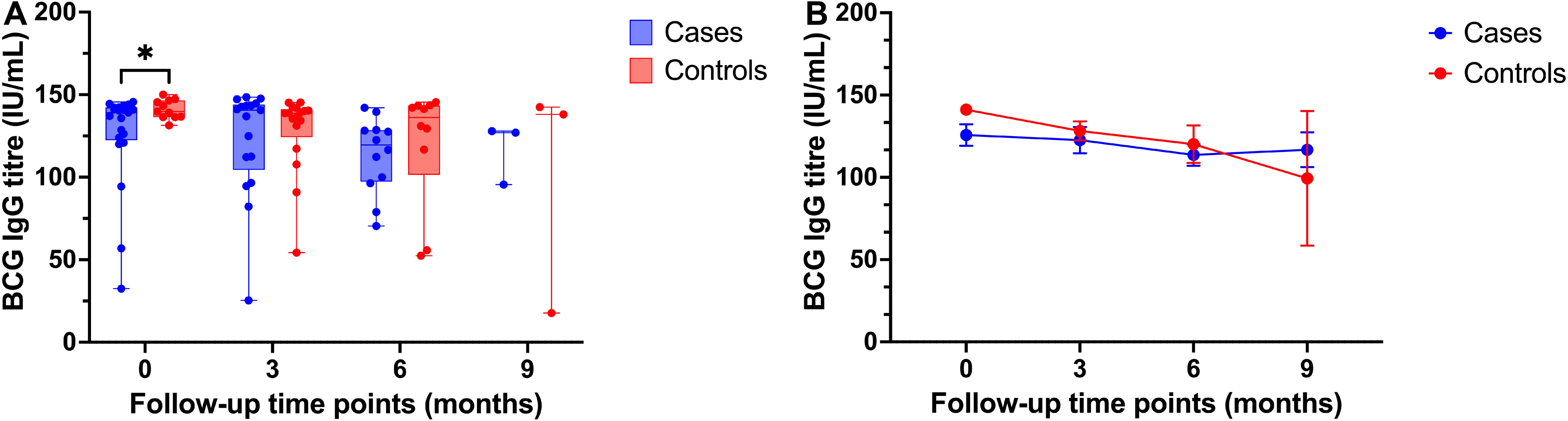
**(A and B).** Plasma was used to perform ELISA using the Human BCG antibody ELISA kit. Two replicates of each sample were used for both case and control samples. The number of samples (n) for cases and controls at V0 was 19/15; at V1, n= 15/15; at V2, n=12/8; and at V3, n=8/3 respectively. Statistical differences were determined using mixed effects analysis and Tukey’s multiple comparison tests. Statistical significance was determined at p<0.05 and a 95% confidence level. * Implies a p-value less than 0.05.

**Table 2:**
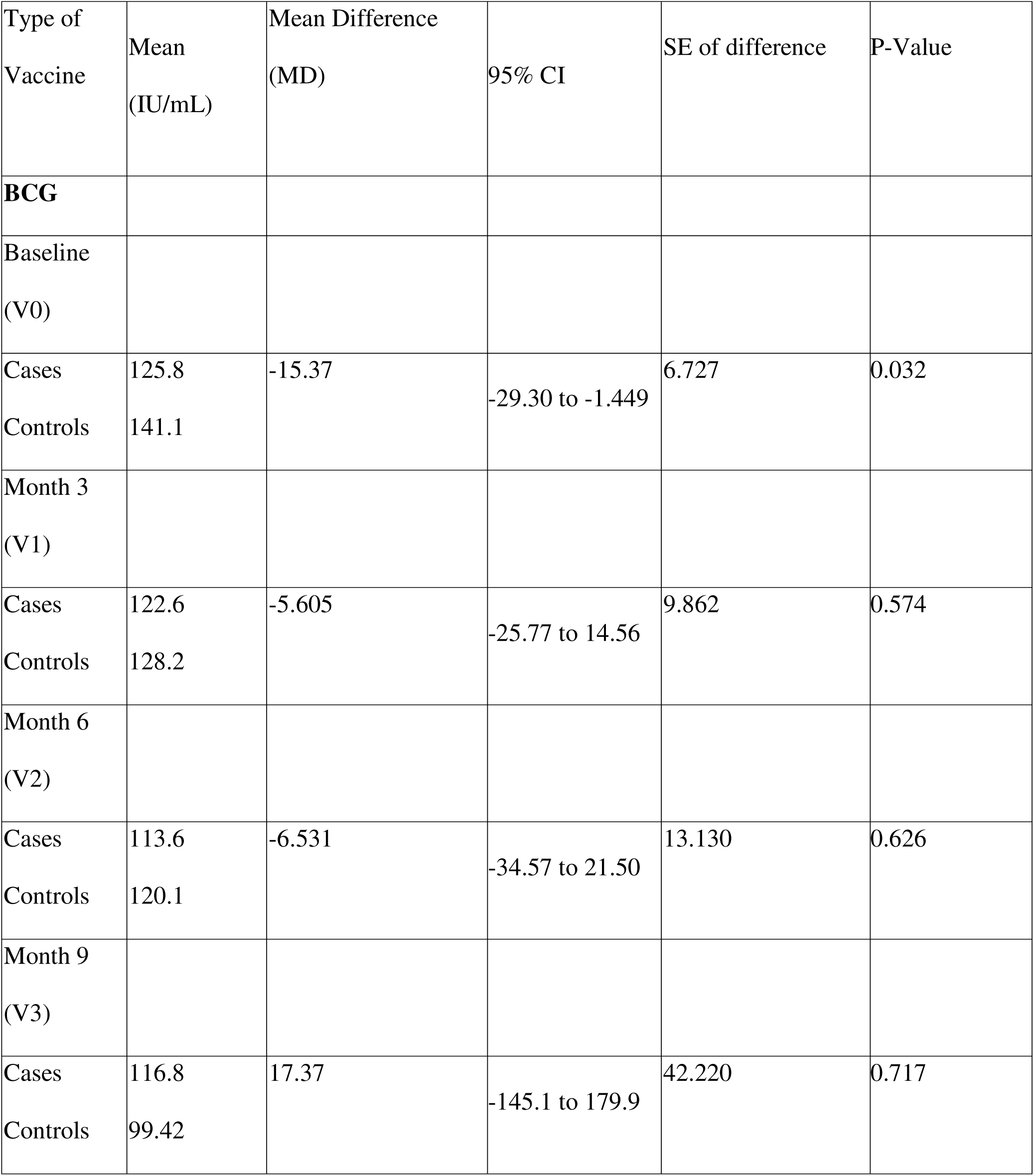
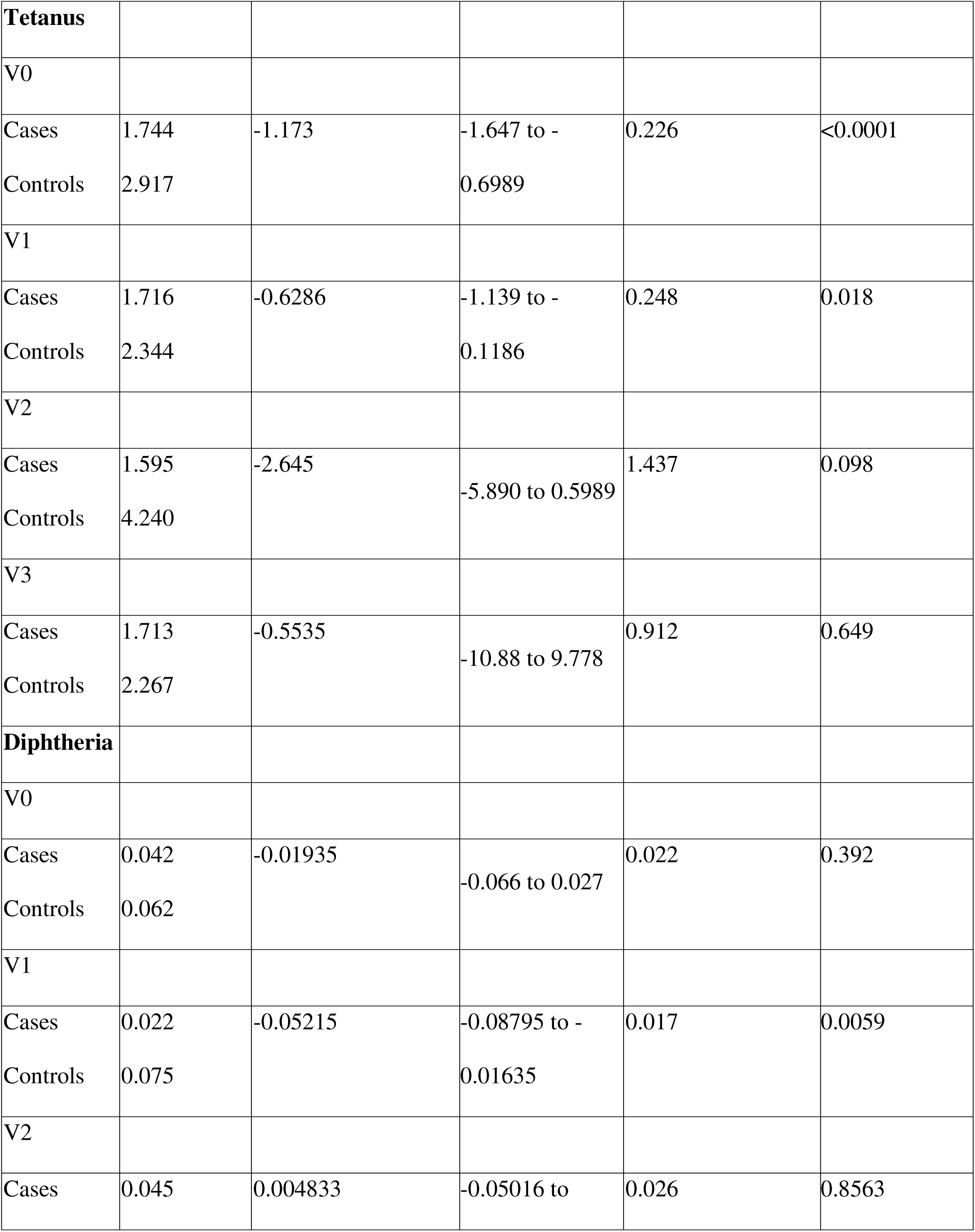
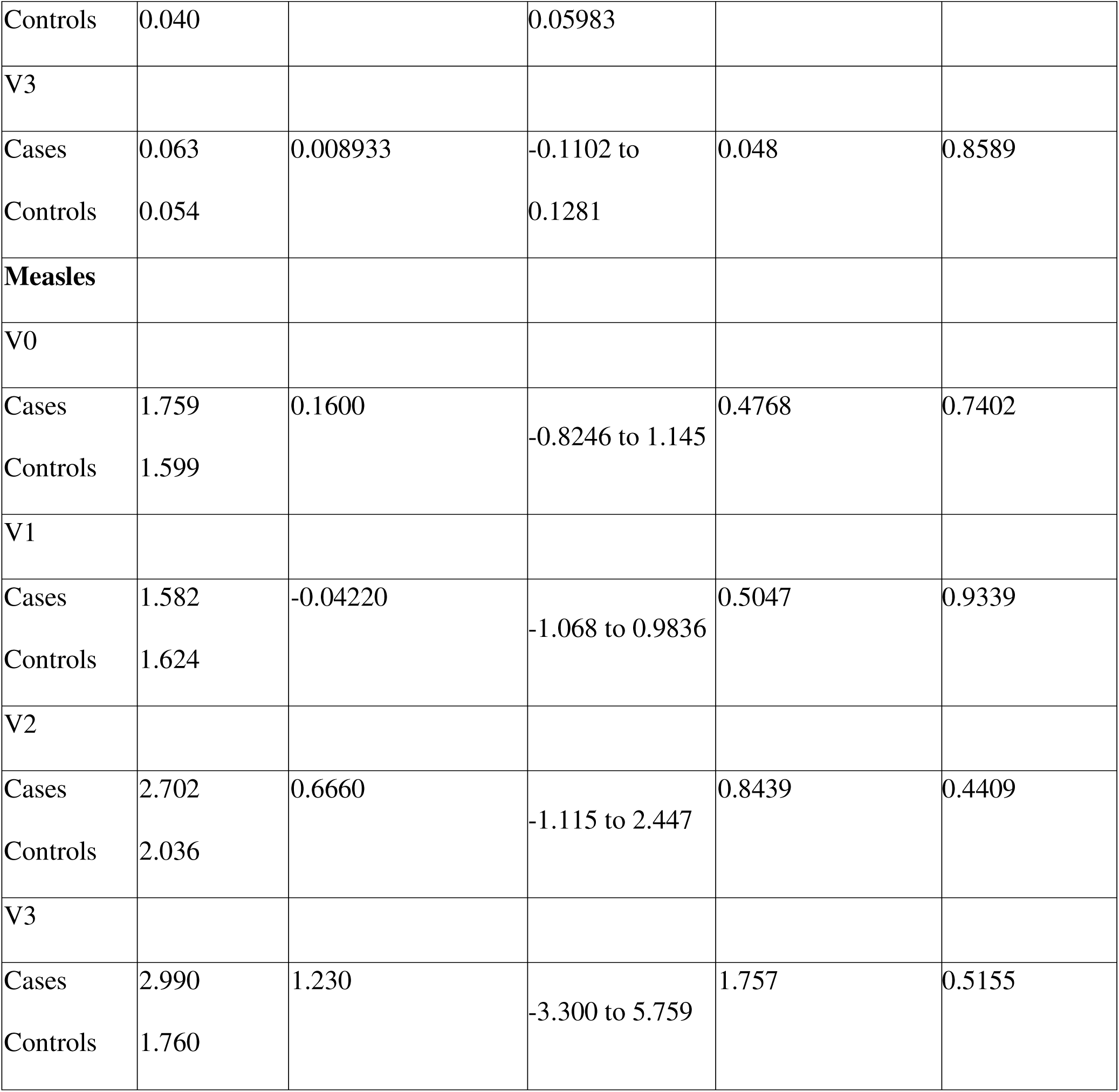
Vaccine responses among cases and control at the different follow-up time points.

### IgG vaccine responses to other vaccines

#### Tetanus-specific IgG responses

Baby cases had slightly lower IgG responses to tetanus at baseline compared to the controls (p<0.0001) and this was followed by a significant decrease at 3 months (p=0.018). The mean (IU/mL), mean difference (MD), 95% confidence interval (CI), P-value, and Standard Error (SE) are shown in Table 2. The trends showed a steep reduction in the responses among controls; however, the tetanus titres among cases increased gently up to 9 months, slightly surpassing the controls observed to fairly rebound at 9 months, as shown in Figure 2(A and B).

**Figure 2.**
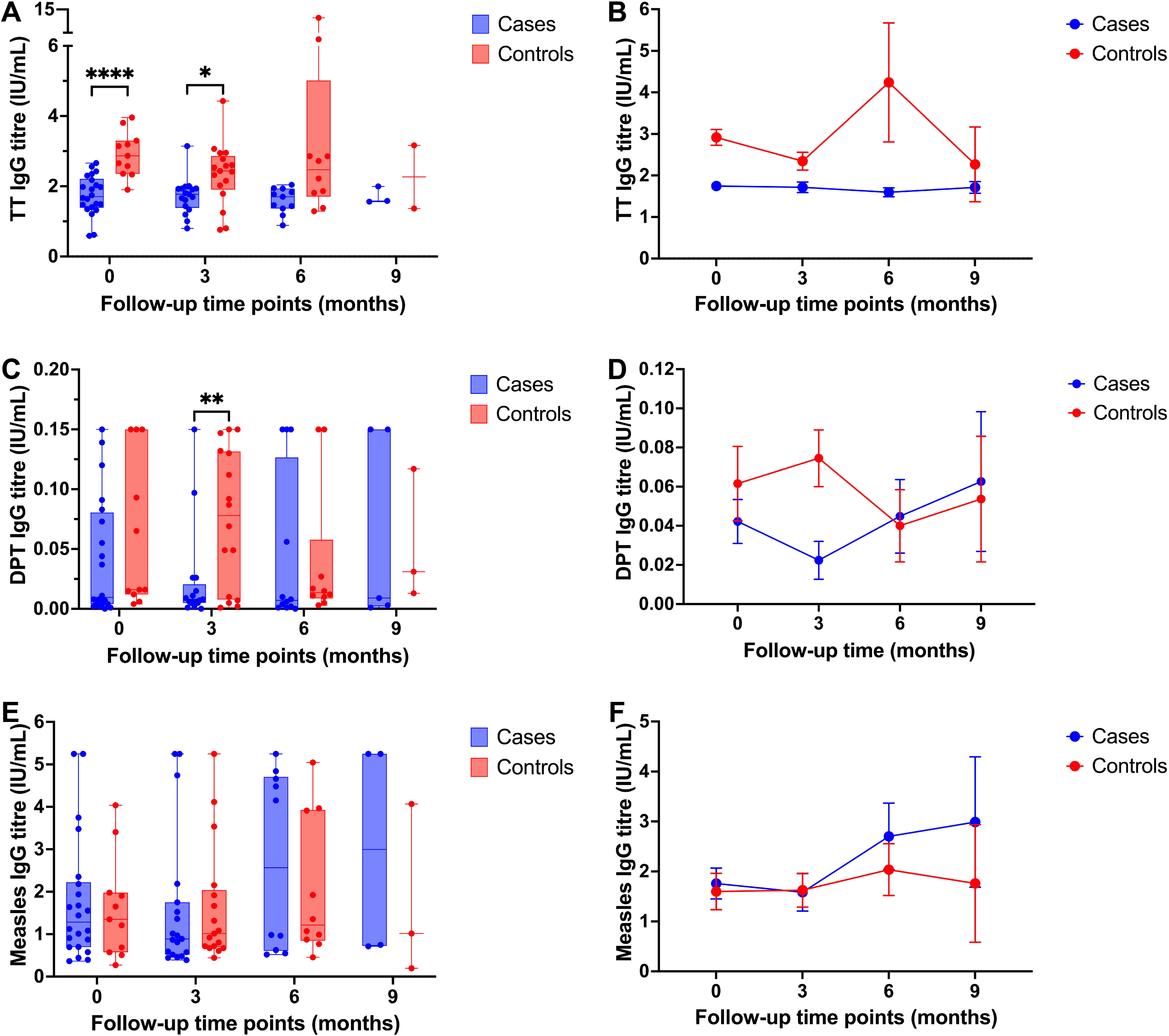
**(A and B).** Stored plasma was used to perform ELISA using the Human Tetanus Toxoid Antibody IgG (TT-IgG) ELISA Kit. The number of cases versus controls used at each visit was V0=18/17, V1=15/11, V2=11/10, and V3=9/4, respectively. Statistical differences were determined using mixed effects analysis and Tukey’s multiple comparison tests. Statistical significance was determined at p<0.05 and a 95% confidence level. * and **** imply a p-value less than 0.05 and 0.0001, respectively. **(C and D).** The plasma was then used to perform ELISA using the Corynebacterium diphtheriae toxin IgG ELISA kit. The number of cases versus controls at V0=20/14, V1=15/15, V2=13/7, and V3=8/4, respectively. Statistical differences were determined using mixed effects analysis and Tukey’s multiple comparison tests. Statistical significance was determined at p<0.05 and a 95% confidence level. ** indicate a p-value less than 0.01. **(E and F).** Plasma was used to perform ELISA using the Human Measles virus IgG antibody (MV-Ab-IgG) ELISA Kit. The number of cases versus controls used at V0=20/16, V1=18/12, V2=11/9, and V3=8/5, respectively. Statistical differences were determined using mixed effects analysis and Tukey’s multiple comparison tests. Statistical significance was determined at p<0.05 and a 95% confidence level.

#### Diphtheria-specific IgG responses

Cases had strongly reduced IgG responses at 3 months of follow-up (p=0.0059) compared to controls. The mean (IU/mL), mean difference (MD) 95% confidence interval (CI), P-value, and Standard Error (SE) are shown in Table 2. A sharp increase followed this in titres, which peaked at 6 months and then dropped at 9 months for controls. On the other hand, the responses for cases are steadily maintained from baseline up to the end of follow-up, as shown in Figure 2(C and D). At months 6 and 9, the p values were p=0.8563 and 0.8589, respectively. No difference was observed in the response to diphtheria at baseline (p=0.392).

#### Measles-specific IgG responses

The trends showed that cases had a slightly higher response at all time points than controls, with the highest response shown at month 9. On the contrary, controls show waning titres between months 6 and 9. At baseline, 3, 6, and 9 months, the p values were: p=0.7402/0.9339/0.4409/0.5155, respectively. These differences were, however, not statistically supported, as shown by Figure 2(E and F). The mean (IU/mL), mean difference (MD) 95% confidence interval (CI), P-value, and Standard Error (SE) are shown in Table 2.

#### Impact of birth weight on vaccine responses

In this study, we further grouped cases and controls as either normal or low weights (kgs) based on the World Health Organisation’s guidelines, and babies with a weight lower than 2.5 kgs were considered low weight. Normal weight ranged between 2.5 and 4.0 kgs. BCG and tetanus-specific IgG titres for normal weight were lower among cases than controls. However, there was no difference in diphtheria and measles vaccine responses between low and normal-weight babies, as shown in Figure 3 (A, B, C, and D). A comparison of the vaccine responses to BCG, tetanus, diphtheria, and measles vaccines and normal birthweight revealed that cases with normal weight had low responses to BCG and tetanus vaccines. No differences were observed among low-weight babies and measles and diphtheria responses. For normal weight (cases versus controls), BCG, p=0.007; tetanus, p=0.004; DPT, p=0.093; measles, p=0.055. The regression analysis also revealed that weight slightly affected BCG responses (p=0.06); however, this was not statistically supported.

**Figure 3.**
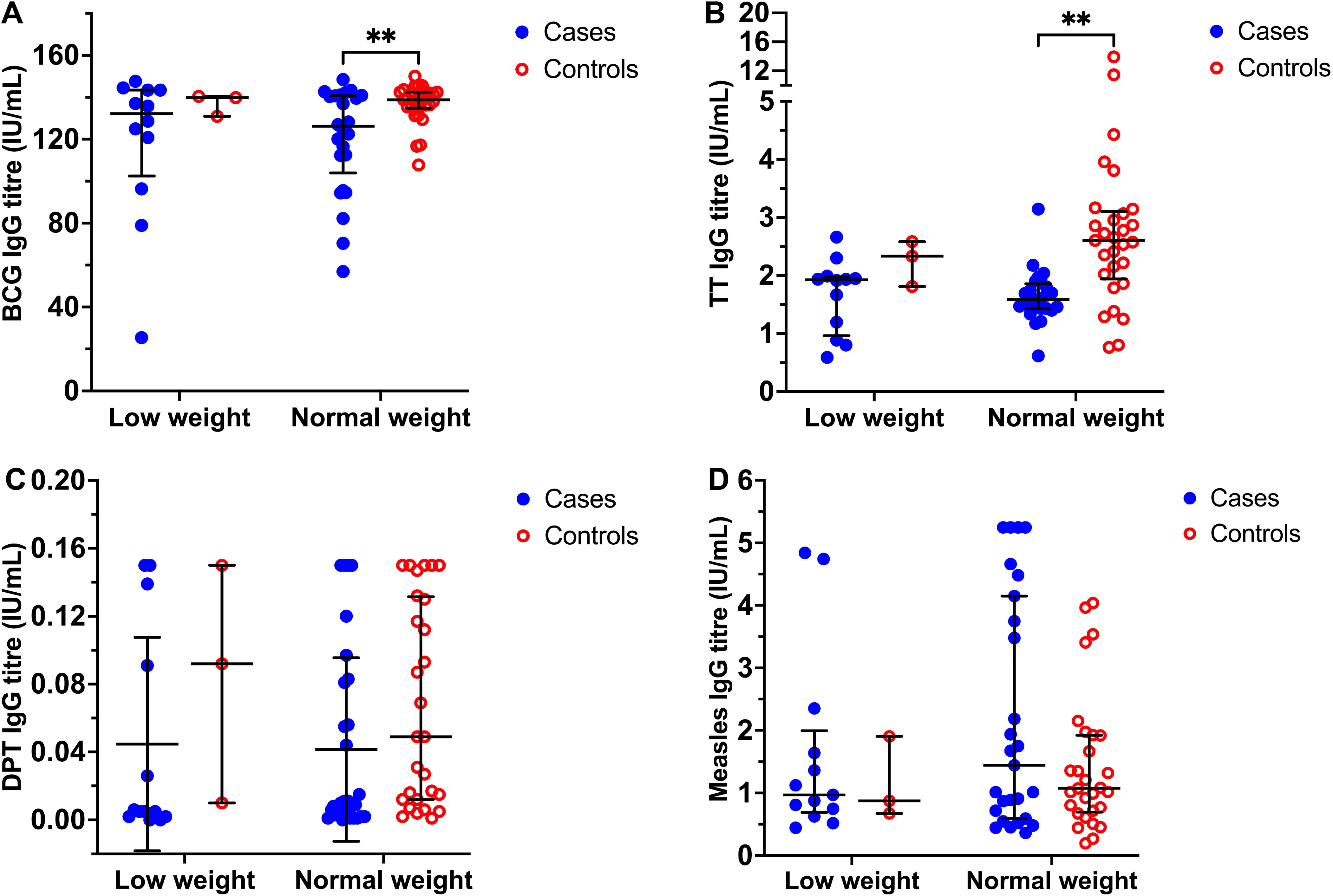
**(A, B, C, D).** This figure shows a sub-analysis of IgG vaccine-specific responses for babies of low and normal weights. Mann-Whitney U test was used to determine the statistical differences. Data were expressed as median and interquartile ranges. ** indicate a p-value less than 0.01.

## Discussion

Pregnancy modulates the immune response towards Th2-type response (12) increasing susceptibility to intracellular pathogens, including TB (13). This study demonstrated that babies born to mothers with ATB have reduced BCG-specific IgG titres at baseline. Maternal tuberculosis disease potentially leads to the transfer of tuberculosis antigens to the foetus, thereby resulting in an altered priming of the immune response (14). The observed reduced BCG responses at baseline could, therefore, result in anergy induced by maternal antibodies; however, this remains to be explored. The responses remained relatively stable over time. The results further show that cases had slightly reduced responses at six months for BCG responses, unlike for Measles. These results contradict what Edwards, 2015 (15) reported: measles vaccine responses were reduced at six months, possibly due to passive maternal antibodies. On the other hand, Nabunya et., 2020 reported that about 36% of infants are not breast fed beyond six months, in Uganda (16). This implies that the transfer of the antibodies is reduced. Our results suggest the insufficiency in responses to the BCG vaccine due to TB exposure. These findings show that maternal TB may impair the infants’ immune responses to the BCG vaccine. We suggest that careful attention is taken to ensure that the infants exposed to TB can develop protective immunity following vaccination to protect infants from early-life infections and deaths from active TB.

In addition to BCG, the study reported a reduction in Diphtheria-specific IgG-specific titres among cases at three months. Tetanus-specific IgG-specific vaccine responses were reduced at baseline and three months among cases compared to the controls. This data highlights that maternal TB disease indeed influences responses to vaccines among infants, and care should be taken to ensure that these exposed infants achieve protective immunity following vaccination. This will be aimed towards averting early-life infections and, consequently, potential infant deaths attributable to active TB. DPT and measles responses are seen to rebound after three months, shooting higher among cases than the controls at 6 and 9 months; however, the differences in fluctuations are not significant. On the other hand, Tetanus responses for cases are relatively stable over time, indicating minimal responsiveness to the vaccine.

The observed decrease in the IgG-specific diphtheria vaccine responses is consistent with findings from similar studies. Abu-Raya et al., 2021 found lower IgG levels in children whose mothers had received the Tdap vaccine (17). Due to the impact of TB in pregnancy, including the low birth weight of babies, vaccination tends to be delayed. When administered, low vaccine responses are more likely to occur (18) predisposing infants to developing vaccine-preventable diseases and experiencing severe or fatal disease outcomes. It has also been demonstrated that maternal *Mtb* infection during pregnancy raises levels of immune-regulatory cytokines (19), which inhibit the infant’s immunological response to vaccinations.

The current study also reports decreased Tetanus-specific IgG vaccine titres among cases. Because the vaccine is administered at six weeks (20), there could have been delayed priming of B cells. In addition, since pregnant women are vaccinated with Tetanus toxoid (21), the maternal IgG that crosses the placenta is likely to induce anergy to the infant’s immune system, thereby causing a dampened immune response. These findings highlight the need for improved vaccine strategies for infants exposed to TB and a better understanding of vaccine failures in certain populations. The effects of TB exposure are also seen with other vaccines like Diphtheria and Tetanus. With vaccine failure in a certain group of people, efforts need to be taken to identify more shortfalls of the existing vaccines and pave the way for effective vaccines possibly tailored to populations. This will enormously contribute to the End TB strategy.

This study further highlights the impact of maternal TB on the birth outcomes of babies, especially low birth weight. Similar results were reported elsewhere (22, 23), affirming this outcome. Vaccination delays associated with low birth weight further hamper the health outcomes of these babies, contributing to lower responses to vaccines and increased vulnerability to infections (24). We postulate that the effects of TB exposure are not only observed with low birth weight. Therefore, when assessing for TB exposure aftereffects, babies with normal weight should not be overlooked. Moreover, the regression analysis revealed that the weight of the babies slightly affected BCG-specific IgG responses, as shown in supplementary figure S1A. Our results also revealed that the mothers to our cases had low haemoglobin concentrations compared to the mothers of the controls. Low haemoglobin during pregnancy is associated with poor neonatal outcomes not limited to preterm births and very low birthweights (25). However, the babies’ responses were not affected by their haemoglobin concentrations following the regression analysis, as shown in Supplementary Figure S1B. These findings show an increased risk for these mothers, especially resulting in preterm babies and low-weight babies.

## Strengths and limitations

In this study, baby case samples were matched with controls regarding age and gender to control for confounder biases. We report that the study had some limitations. The ATB yield was low, which affected the number of participants studied. However, we were able to demonstrate a statistically supported difference. Some of our participants were enrolled during puerperium, so the babies received the initial vaccines before enrolment. This limited our understanding of the initial status of these babies before vaccination. Also, data on mothers’ vaccination status against tetanus during pregnancy was not collected, and we can only anticipate that since it is part of routine antenatal care, they all received this vaccine. We anticipate that this could have further dampened immune responses to the vaccine among cases.

## Conclusion

Babies born to mothers with active TB disease: (i) Had low birth weights, (ii) Presented with reduced vaccine-specific IgG responses for BCG and the other infant vaccines in general but markedly at baseline. This could be resulting from persistent *Mycobacterial tuberculosis* antigenemia. We speculate a similar scenario to the *DoHAD* theory. We propose a larger prospective study with longer follow-ups to explore outcomes of vaccination and TB disease risk. This would also answer whether the suppressed response to the vaccine among exposed infants is due to other factors such as the participant’s household or environment and nutrition, in addition to revealing whether the responses improve or completely diminish over time. A study on determining the quality of antibodies from active TB mothers, which would look at antibody affinity and avidity to compare these between cases and controls, is necessary. Finally, looking at the transcriptomic changes in the blood of these infants exposed to TB would be insightful. This would inform some of the highly expressed biomarkers potentially used to diagnose childhood TB.

## Supporting information

Supplemental Figure 1: Haemoglobin concentration versus baby responses

Study protocols and Supplemental Figure 1 legend

## Author contributions

DS performed laboratory experiments, analysed data, and drafted the manuscript. IAB, AN, SC, RN, FB, JBB, PS, APK, DK, and NR participated in the concept development and reviewed the manuscript. PS developed the experimental protocols that followed the experiment and offered technical laboratory training to perform the experiments. IAB sourced funding for the research.

## Data availability

The authors will make raw data that guided these conclusions available upon request.

## Acknowledgement

A vote of special thanks goes to the Tuberculosis and Comorbidities Research Consortium’s field team for data collection. We thank the study participants and the health facility administrators for their engagement in this study. This work was supported by the Crick African Network, which receives its funding from the UK’s Global Challenges Research Fund (MR/P028071/1), and by the Francis Crick Institute, which receives its core funding from Cancer Research UK (FC1001647), the UK Medical Research Council (FC1001647), and the Wellcome Trust (FC1001647). We also acknowledge HEPI-SHSS for offering a research contribution to the laboratory work. This work was presented at conferences and an abstract of this work was published as a conference paper in BMJ, by the EDCTP Forum, 2023.

