## Supplementary figures and images for "Reduced *Bacille Calmette-Guérin*-specific IgG titres among babies born to mothers with Active Tuberculosis Disease in Uganda"

### Supplemental Figure 1: Haemoglobin concentration versus baby responses

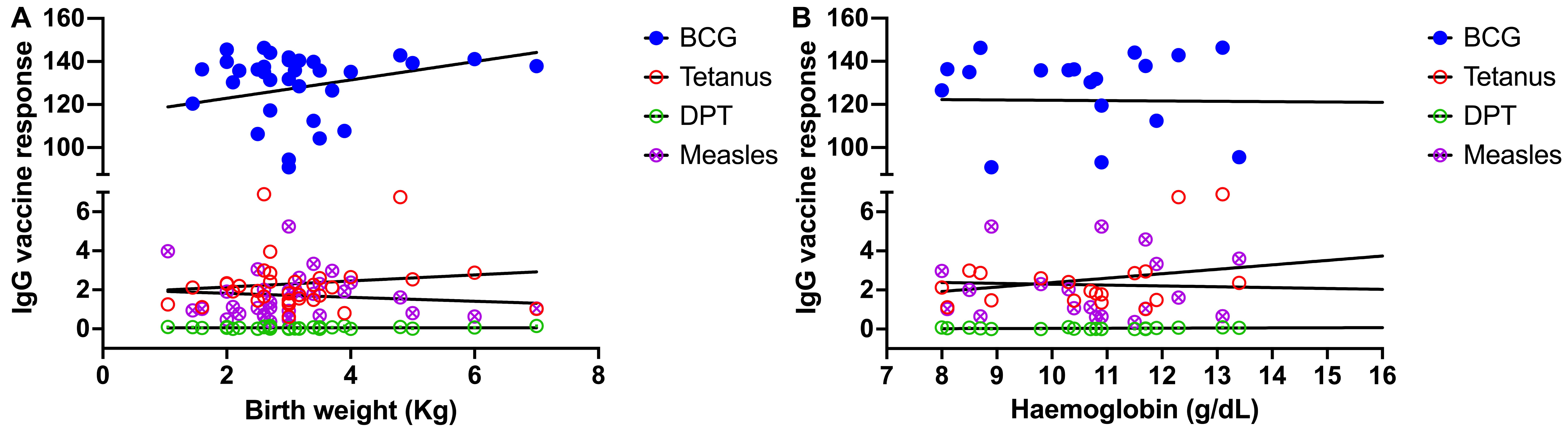
