## Supplementary material for "Reduced *Bacille Calmette-Guérin*-specific IgG titres among babies born to mothers with Active Tuberculosis Disease in Uganda": Study protocols and Supplemental Figure 1 legend

**Supplementary information**

### 1. Procedure for Human BCG antibody (IgG) ELISA

The microtiter plates provided in this kit were pre-coated with *Mtb* antigen. Optimization was performed and seven cryo vials were used. The first vial had an undiluted sample, and we performed 1:20 dilution for the second vial, 1:40 for the third vial, 1:80 for the fourth vial, 1:160 for the fifth vial, 1:320 for the sixth vial, and finally 1:640 for the seventh vial. Suitable results were observed in the undiluted sample. A blank well was set first in duplicate and then three Negative Control wells and two Positive Control wells were selected. 100μl of Negative Control or Positive Control were added per well and this was followed by 100μl of undiluted sample to the rest of the wells.

Plates were sealed and incubated for 30 minutes at 37°C and thereafter washed with 300 microliters of 1X wash buffer five times, using an automated well wash, and blotted over a paper towel to ensure complete drying. 100μl of HRP-conjugate was added to each well using a multichannel pipette, except for the blanks. Plates were then sealed and incubated for 30 minutes at 37°C and thereafter washed again five times with 300 microliters of the 1X wash buffer, using an automated well wash. 50μl of Substrate A and 50μl Substrate B were added to each of the wells and the plates were incubated for 10 minutes at 37°C, away from light, and covered with aluminum foil. 50μl of Stop Solution was added to each of the wells using a multichannel pipette, and plates were shaken on a plate shaker to ensure uniform mixing and thereafter transferred to the ELISA reader (Synergy LX multimode reader). Using the Gen 5 software version 2.71.2, the wavelength was set to 450 nm, and considering the blank wells were taken as zero, the optical density of each of the wells was determined.

### 2. Procedure for Corynebacterium diphtheriae toxin IgG ELISA

The principle of this assay was: that microtiter plates were pre-coated with specific antigens to bind corresponding antibodies of the sample. Optimization was performed and seven cryo vials were used. The first vial had an undiluted sample, and we performed 1:20 dilution for the second vial, 1:40 for the third vial, 1:80 for the fourth vial, 1:160 for the fifth vial, 1:320 for the sixth vial, and finally 1:640 for the seventh vial. Suitable results were observed in 1:640 sample dilution. 100 microliters of standards and diluted samples were dispensed into their selected respective wells and well A1 was left out for the Substrate Blank. Wells were completely sealed with aluminum foil and incubated in an incubator at 37°C. Following incubation, the contents of the plate wells were aspirated using a multichannel pipette, ensuring to change tips after every column. Plates were then washed three times with 300 microliters of 1X Washing Buffer, using an automated well wash, at an interval of 10 seconds and thereafter blotted on a paper towel to completely dry. 100 microliters of Conjugate were dispensed into all wells except for the Substrate Blank well A1 and incubated for 30 min at room temperature, away from light. Plates were then washed as described above and 100 microliters of TMB Substrate Solution were added into all wells using a multichannel pipette. This was followed by a 15-minute incubation at room temperature (20 to 25 °C) in the dark and thereafter, 100μl of stop solution was dispensed into all wells in the same order and at the same rate as for the TMB Substrate Solution, and a color change from blue to yellow was observed. The absorbance at 450/620 nm was measured using the Gen 5 software version 2.71.2 for the ELISA reader (Synergy LX multimode reader).

### 3. Procedure for Human Tetanus Toxoid Antibody IgG (TT-IgG) ELISA

For this Quantitative Sandwich ELISA kit, Tetanus antigen was bound on the surface of the micro titer strips. Optimization was performed and seven cryo vials were used. The first vial had an undiluted sample, and we performed 1:20 dilution for the second vial, 1:40 for the third vial, 1:80 for the fourth vial, 1:160 for the fifth vial, 1:320 for the sixth vial, and finally 1:640 for the seventh vial. Suitable results were observed in 1:640 sample dilution. Standard, sample, and blank wells were set and 50 μl of reconstituted standards (S1, S2, S3, S4, S5, S6) were added to their corresponding Standard wells. This was followed by the addition of 50 μl of diluted sample to every sample well. 100 μl HRP-Conjugate Reagent was added to every well except blank wells and plates were sealed with aluminum foil and incubated for 60 minutes at 37°C. Wells were then washed with 300 μl 1X wash buffer using an automated well wash, at intervals of 10 seconds of soak time, four times. 50 μl of both Chromogen Solutions A and B were added to each well, plates were shaken using a plate shaker and then incubated for 15 minutes at 37°C in the dark. 50 μl of stop solution was added to every well, and optical density (O.D) at 450 nm was obtained in the Gen 5 software version 2.71.2 of the ELISA reader (Synergy LX multimode reader).

### 4. Procedure for Human Measles virus IgG antibody (MV-Ab-IgG) ELISA

The microtiter plate provided in this kit was pre-coated with the measles virus. Optimization was performed and seven cryo vials were used. The first vial had an undiluted sample, and we performed 1:20 dilution for the second vial, 1:40 for the third vial, 1:80 for the fourth vial, 1:160 for the fifth vial, 1:320 for the sixth vial, and finally 1:640 for the seventh vial. Suitable results were observed in the undiluted sample. Blank wells were set and no solution was added in them. 100 μl of Negative Control, Positive Control and undiluted samples were added in the selected wells and plate was incubated for 30 minutes at 37°C. Contents of the wells were then aspirated and 250 μl of 1X wash buffer was used to wash the plates with the aid of the automated well wash, at soak intervals of 10 seconds and later blotted on a paper towel to ensure complete drying. 100 μl of HRP-conjugate was added to each well except the blank wells and plates were covered with adhesive films and also sealed in aluminum foil and thereafter incubated for 30 minutes at 37°C. Washing was repeated using the automated well wash and 50 μl of substrates A and B were added to all wells, the plate was shaken and incubated for 10 minutes at 37°C, in the dark. 50 μl of Stop Solution was added to each well and the plate was shaken gently. Plates were then transferred to the ELISA reader (Synergy LX multimode reader) to determine optical densities at 450nm and 630 nm wavelengths, using the Gen 5 software version 2.71.2.

**Legend for supplementary figure S1.**

**Figure S1 (A and B).**

A simple linear regression was performed to determine the effect of birth weight (S1A) and hemoglobin (S1B) after which the Spearman rank coefficient test was done. A positive correlation was observed between birth weight and BCG responses however this was not statistically supported. A p-value of less than 0.05 was considered statistically significant.
